# Stimulant medication use and apparent cortical thickness development in attention-deficit/hyperactivity disorder: a prospective longitudinal study

**DOI:** 10.1101/2023.07.28.23293202

**Authors:** Zarah van der Pal, Kristine B Walhovd, Inge K Amlien, Carlijn Jamila Guichelaar, Antonia Kaiser, Marco A Bottelier, Hilde M Geurts, Liesbeth Reneman, Anouk Schrantee

## Abstract

**Background:** Stimulant medication is commonly prescribed as treatment for attention-deficit/hyperactivity disorder (ADHD). While we previously found that short-term stimulant-treatment influences apparent cortical thickness development in an age-dependent manner, it remains unknown whether these effects persist throughout development into adulthood.

**Purpose:** Investigate the long-term age-dependent effects of stimulant medication on apparent cortical thickness development in adolescents and adults previously diagnosed with ADHD.

**Methods:** This prospective study included the baseline and 4-year follow-up assessment of the “effects of Psychotropic drugs On the Developing brain-MPH” (“ePOD-MPH”) project, conducted between June-1-2011 and December-28-2019. The analyses were pre-registered (https://doi.org/10.17605/OSF.IO/32BHF). T1-weighted MR scans were obtained from male adolescents and adults, and cortical thickness was estimated for predefined regions of interest (ROIs) using Freesurfer. We determined medication use and assessed symptoms of ADHD, anxiety, and depression at both time points. Linear mixed models were constructed to assess main effects and interactions of stimulant medication use, time, and age group on regional apparent cortical thickness.

**Results:** A total of 32 male adolescents (aged mean±SD, 11.2±0.9 at baseline) and 24 men (aged mean±SD, 29.9±5.0 at baseline) were included that previously participated in the ePOD-MPH project. We found no evidence for long-term effects of stimulant medication use on ROI apparent cortical thickness. As expected, we did find age-by-time interaction effects in all ROIs (left prefrontal ROI: P=.002, right medial and posterior ROIs: P<.001), reflecting reductions in apparent cortical thickness in adolescents. Additionally, ADHD symptom severity (adolescents: P<.001, adults: P=.001) and anxiety symptoms (adolescents: P=0.03) were reduced, and lower change in ADHD symptoms was associated with higher medication use in adults (P=0.001).

**Conclusion:** We found no evidence for long-term effects of stimulant-treatment for ADHD on apparent cortical thickness development in adolescents and adults. The identified age-dependent differences in apparent cortical thickness development are consistent with existing literature on typical cortical development.

**Summary statement:** This prospective longitudinal study in male adolescents and adults found that stimulant medication does not modulate long-term apparent cortical thickness development.

**Key results:**

- In this prospective longitudinal structural MRI study of 32 male adolescents and 24 men previously diagnosed with attention-deficit/hyperactivity disorder (ADHD), we found no evidence for long-term effects of stimulant medication on apparent cortical thickness development.
- We identified age-dependent patterns of cortical development, with reductions in apparent cortical thickness in adolescents only (P<.001 in all regions of interest).

## Introduction

Stimulant medication, such as methylphenidate-(MPH) and dexamphetamine-based formulations, is commonly prescribed as treatment for ADHD, a prevalent neurodevelopmental disorder characterised by age-inappropriate levels of inattentive, hyperactive and impulsive behaviour [1,2]. Stimulant medication has been shown to be highly effective in alleviating core ADHD symptoms of hyperactivity and inattention, as well as ancillary symptoms such as emotional dysregulation [3,4]. Although children and adolescents often receive stimulant-treatment for extended periods of time, possible long-lasting effects of extended stimulant-treatment on cortical development of the brain remain unclear.

Cortical morphology undergoes continuous development throughout the lifetime, with magnetic resonance imaging (MRI) studies reporting rapid reductions in apparent cortical thickness (i.e., cortical thinning) during adolescence and continued cortical thinning at a slower rate throughout adulthood [5,6]. In contrast, changes in cortical surface area predominantly occur during childhood and early adolescence [7,8]. Previous studies investigating cortical maturation in individuals with ADHD using MRI suggest that children and adolescents with ADHD ‘lag behind’ typically developing peers in development of grey matter volume and cortical thickness, particularly in prefrontal regions [9]. Moreover, alterations in cortical thickness, surface area and grey matter volume have been negatively associated with clinical outcomes such as ADHD symptom severity and depressive symptoms [10,11]. Of note, apparent changes in cortical thickness during development may in part result from other factors such as increased myelination, which impacts MR contrast and the grey-white matter boundaries used for cortical thickness estimation [12].

However, studies investigating the effect of stimulant medication on brain morphology are less clear and yielded inconsistent results. For instance, a longitudinal study reported more rapid cortical thinning in ADHD participants *off* stimulant medication, compared with ADHD participants *on* stimulant medication and typically developing peers [13]. Furthermore, a voxel-based morphometry meta-analysis found that stimulant medication use was associated with higher (i.e., more “normalised”) grey matter volume in the right caudate [14]. In contrast, two large-scale studies using cross-sectional data in adolescents and adults with ADHD identified no associations between various stimulant-treatment parameters and cortical thickness [15,16]. Notably, prior research mostly examined children, adolescents, or adults separately, without considering potential age-related effects of stimulant-treatment on cortical development. Such age-dependent effects of stimulant medication are supported by animal studies, suggesting that stimulant medication use during early adolescence has lasting effects on brain development (“the neurochemical imprinting hypothesis”) [17,18]. Moreover, we previously found that 4-month MPH-treatment resulted in less rapid cortical thinning in children with ADHD, but not in adults or placebo groups [19]. Thus, (preclinical) findings so far suggest that the short-term effects of stimulant-treatment on apparent cortical thickness development are modulated by age. However, it remains unclear whether these effects last throughout development into adulthood.

We hypothesised that stimulant medication use would induce persistent (long-term) age-dependent effects on regional cortical thickness. Specifically, we hypothesised that higher stimulant exposure would be associated with less rapid regional cortical thinning in adolescents, but not in adults with ADHD. Therefore, the present study aimed to investigate whether stimulant medication modulates regional cortical thickness development in a 4-year naturalistic follow-up of the children and adults previously diagnosed with ADHD.

## Methods

### Study design

This study is part of the prospective “effects of Psychotropic drugs On the Developing brain - MPH” (ePOD-MPH) project. The initial ePOD-MPH randomised controlled trial (RCT) was a 16-week double-blind, randomised, placebo-controlled, multicentre trial of MPH-treatment conducted between June 1 2011 and June 15 2015, with a blinded endpoint evaluation in stimulant treatment-naive participants with ADHD [20]. The present study constitutes the naturalistic 4-year follow-up assessment of the ePOD-MPH RCT, conducted between March 1 2016 and December 28 2019. The ePOD-MPH RCT protocol applied the code of medical ethics and was registered by the Central Committee on Research Involving Human Subjects (an independent registry) on March 24, 2011 (identifier NL34509.000.10) and subsequently at The Netherlands National Trial Register (identifier NTR3103), with enrolment of the first patient on October 13, 2011. The 4-year follow-up assessment was approved by the local medical ethical committee of the Academic Medical Center (NL54972.018.15). All participants and parents or legal representatives of the children provided written informed consent.

This study’s design and analysis plan were pre-registered at the Open Science Foundation registry (https://doi.org/10.17605/OSF.IO/32BHF). For deviations from the pre-registered analysis plan, please see Supplemental Materials.

### Participants

For the initial ePOD-MPH RCT, we included 50 children (aged 10-12 years) and 49 adult (aged 23-30 years) male outpatients diagnosed with ADHD (all subtypes). ADHD diagnosis, as defined in the Diagnostic and Statistical Manual of Mental Disorders, Fourth Edition (DSM-IV [1]), was determined by an experienced psychiatrist using a structured interview (Diagnostic Interview Schedule for Children fourth edition, DISC-IV [21]) in children or in parents and the Diagnostic Interview for Adult ADHD [22] in adults (for details on recruitment and exclusion criteria, see Supplemental Materials). For the current study, the 4-year follow-up assessment, participants were contacted by phone and/or email to ask if they wanted to participate in the follow-up assessment. Exclusion criteria were contraindications to MRI.

### Medication use

Stimulant medication use per participant was calculated based on medication received during the initial ePOD-MPH RCT and medication use information between the ePOD-MPH RCT and 4-year follow-up assessment obtained from pharmacies. The following medication use variables were calculated: cumulative dose (mg), exposure duration (months), mean daily dose (mg/day), and age at start of medication use (years). Moreover, we determined stimulant medication use at follow-up (yes/no) and stimulant treatment-naivety at follow-up (yes/no) (for details, see Supplemental Materials).

During the ePOD-MPH RCT, the treating physician prescribed the study medication under double-blind clinical guidance (reduction of ADHD symptoms) following Dutch treatment guidelines. Exposure duration was 4 months for participants who received MPH, and 0 months for participants who received placebo.

Stimulant medication use between ePOD-MPH RCT end and 4-year follow-up assessment was calculated based on medication history information obtained from participants’ pharmacies. MPH- and dexamphetamine-based formulations were considered as stimulant medication as treatment for ADHD, and cumulative dose was converted to MPH-equivalents [23].

### Clinical and behavioural assessment

ADHD symptoms were assessed using the Disruptive Behavior Disorder Rating-Scale (DBD-RS, inattention and hyperactivity/impulsivity subscales [24]) in adolescents, and the ADHD-Rating Scale (ADHD-RS [22]) in adults. Symptoms of depression and anxiety were assessed using the Child Depression Inventory (CDI [25]) and the child version of the Screen for Child Anxiety Related Disorders (SCARED [26]) in adolescents, and the Beck Depression Inventory (BDI-II [27]) and the Beck Anxiety Inventory (BAI [28]) in adults.

### MR acquisition and processing

At baseline, MR imaging was performed using a 3T Intera or Achieva MR scanner, and at 4-year follow-up using a 3T Ingenia MR scanner (Philips Medical Systems, Best, The Netherlands). 3D T1-weighted fast-field echo sequences (TR/TE/FA=9.8/4.6ms/8°, voxel size=0.875×0.875×1.2 mm, slices=120, reconstruction matrix=256) were acquired at baseline and 4-year follow-up using an 8-channel receive-only head coil. We used a one-week washout period prior to the follow-up assessment to exclude possible acute effects of medication use (half-life: 2-10 hours).

Vertex-wise cortical thickness was estimated across the brain surface using the FreeSurfer longitudinal processing stream (version 7.1 [29]). Predefined ROIs in the left prefrontal, right medial and right posterior parietal cortices (Figure 3A), were selected based on prior findings of psychostimulant effects on apparent cortical thickness by an observational study as well as the initial ePOD-MPH RCT [13,19] (see Supplemental Material for details). Surface measures were extracted from the individual participants. In line with the initial ePOD-MPH RCT, scans were visually inspected and rated for the presence of motion and were excluded unless rated 1 (no sign of motion) or 2 (minor signs of motion, but no major distortion and acceptable reconstruction). Reconstructed datasets were not edited, to avoid manual interference.

### Statistical analysis

Analyses were performed using R version 4.0.3 (R Development Core Team, 2011). To identify potential covariates to include in the statistical analysis, we assessed correlations between medication use variables and other (clinical) outcome measures (Supplemental Materials).

For the confirmatory ROI analysis, linear mixed models (LMMs) were constructed to assess main effects and interactions of stimulant medication use, time (baseline, follow-up) and age group (adolescents, adults) on regional cortical thickness. Separate models were used for each ROI and medication use variable (cumulative dose, exposure duration). Medication use variables were included as 1) continuous, and 2) grouped (none/low vs. high; median split) variables. In addition, to assess whether the effects of the initial ePOD-MPH RCT were still present at 4-year follow-up, we constructed LMMs to assess main effects and interactions of ePOD-MPH RCT treatment group (MPH, placebo), time (baseline, follow-up) and age group (adolescents, adults) on regional cortical thickness. Two covariates were included in all models: demeaned age at baseline (per age group; to correct for the larger age range at baseline among adults compared with adolescents) and standardised scan interval (offset from 48 months, corresponding to 4-year follow-up). MR scanner at baseline was included as a covariate, and removed if it did not significantly improve the model. The significance level was set at P<0.05. Additionally, Bayes Factors were calculated comparing the models with medication use to the models without medication use, to determine the strength of the reported evidence (for details, see Supplemental Materials).

Finally, we performed exploratory analyses evaluating relations between regional cortical thickness and clinical outcomes, and assessed associations between stimulant medication and whole-brain cortical thickness and surface area (Supplemental Materials).

## Results

### Participant characteristics

Of the 50 children and 49 adults who participated in the initial ePOD-MPH RCT, 33 children (66% return rate) and 25 adults (50% return rate) were included in the 4-year follow-up assessment (Table 1, Figure 1). Baseline characteristics were comparable for participants that did and did not return for follow-up (see Supplemental Material). Of those that returned at follow-up, six adolescents were scanned using MR scanner 1 (Intera) and the remaining participants were scanned using MR scanner 2 (Achieva) at the baseline assessment. One adolescent and one adult were excluded from analysis due to missing MRI data and undisclosed prior exposure to stimulant medication, respectively. The final sample consisted of 32 adolescents (aged 11.2±0.9 at baseline) and 24 men (aged 29.9±5.0 at baseline). For two adolescents, MRI data from one time point was excluded (one baseline scan, one follow-up scan), due to incorrect/failed segmentation resulting from motion. In contrast to the ePOD-MPH RCT, two participants with structural brain abnormalities (one adolescent with a posterior/cerebellar cyst, one adolescent with a benign tumour in the right frontal lobe) were not excluded from analysis. This decision was made, since these structural abnormalities did not change between baseline and follow-up, and sample size at follow-up was limited. Results were robust when these participants were excluded in a sensitivity analysis (Supplemental Materials).

**Figure 1.**
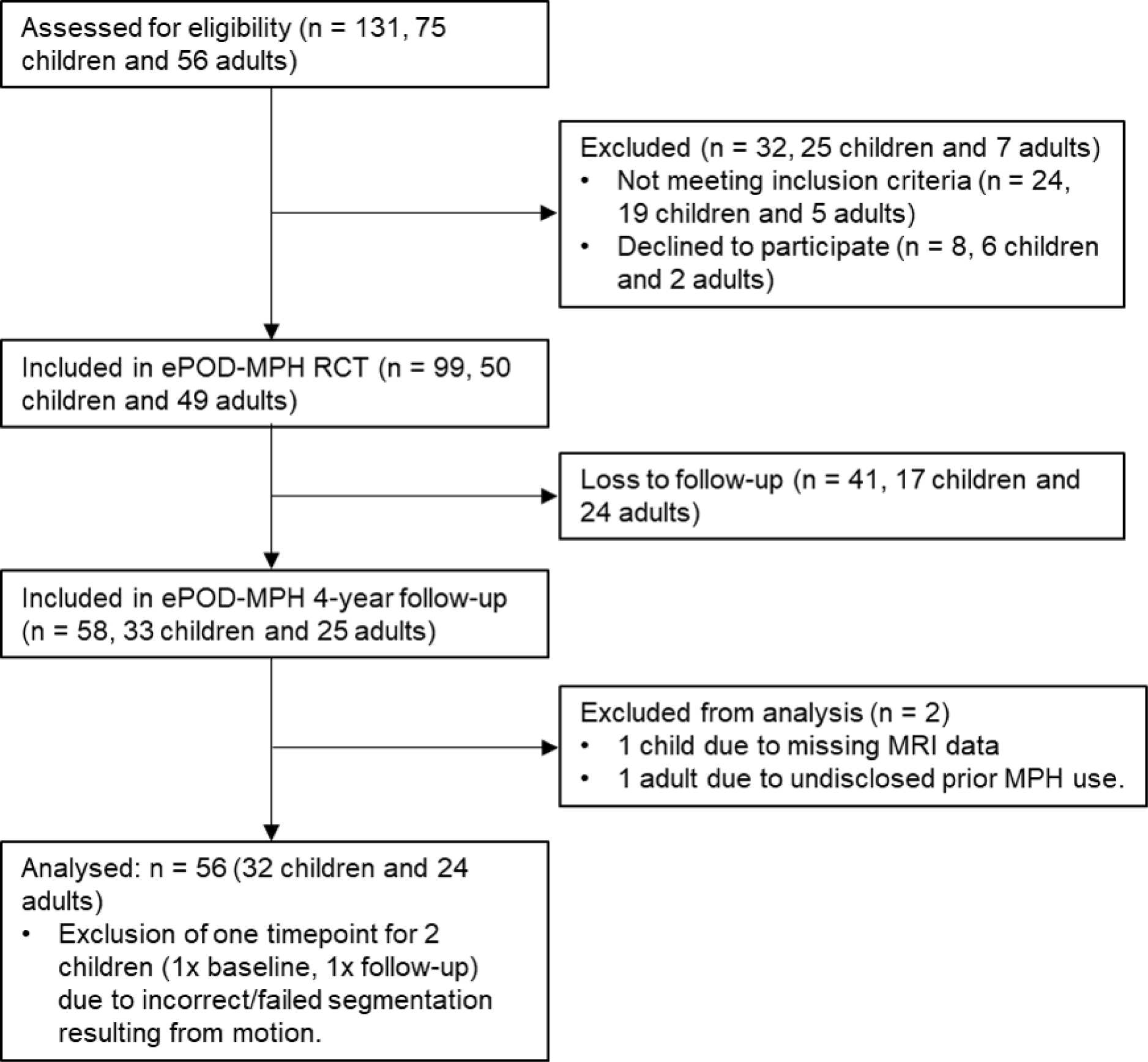
Flow diagram showing participant inclusion process. For consistency, child/adolescent participants are referred to as children throughout the flow diagram. MPH = methylphenidate, RCT = randomised controlled trial.

**Table 1.** Participant characteristics of the study participants. Data are presented as mean (standard deviation), median (interquartile range), fraction (yes/no, MPH/placebo) or percentage (%). All participants were male.

|  | Adolescents<br>n = 32 |  |  | Adults<br>n = 24 |  |  |
| --- | --- | --- | --- | --- | --- | --- |
|  | BL | FU | Statistics<sup>a</sup> | BL | FU | Statistics<sup>a</sup> |
| **Demographics** |  |  |  |  |  |  |
| Age (years, *mean (SD)*) | 11.2 (0.9) | 15.3 (1.4) |  | 29.9 (5.0) | 34.4 (4.6) |  |
| Estimated IQ<sup>b</sup> (*mean (SD)*) | 104.5 (17.7) |  |  | 106.7 (5.0) |  |  |
| Handedness (% *right-handed*) | 93.8 |  |  | 91.7 |  |  |
| **Clinical outcomes (*mean (SD)*)** |  |  |  |  |  |  |
| ADHD-inattentive symptom severity<sup>c</sup> | 22.75 (3.38) | 11.77 (5.20) | t(29) = 11.81, p < .001, BL > FU |  |  |  |
| ADHD-hyperactive/impulsive symptom severity<sup>c</sup> | 15.88 (5.59) | 7.17 (4.41) | t(29) = 10.08, p < .001, BL > FU |  |  |  |
| ADHD symptom severity<sup>d</sup> |  |  |  | 33.13 (9.91) | 24.47 (8.54) | t(18) = 3.75, p = .001, BL > FU |
| Anxiety symptoms<sup>e</sup> | 25.72 (16.75) | 17.52 (13.25) | t(28) = 2.40, p = .02 | 7.13 (6.60) | 4.40 (5.70) | t(19) = 1.51, p = .15 |
| Depressive symptoms<sup>f</sup> | 7.97 (4.40) | 8.61 (4.44) | t(29) = -0.50, p = .62 | 6.21 (4.90) | 5.48 (5.88) | t(20) = 0.28, p = .78 |
| **Medication** |  |  |  |  |  |  |
| Age at start (years, *mean (SD)*) |  | 11.8 (1.4) |  |  | 29.2 (4.0) |  |
| Cumulative dose<sup>g</sup> (mg, *median (IQR)*) |  |  |  |  |  |  |
| total |  | 9455<br>(3533 - 23630) |  |  | 4778<br>(750 - 13228) | W = 456, p = .24 |
| none/low group |  | 3550<br>(2124 - 6342) |  |  | 600<br>(0 - 3795) |  |
| high group |  | 23778<br>(19605 - 30384) |  |  | 14633<br>(6186 - 46179) |  |
| Exposure duration<sup>h</sup> (months, *median (IQR)*) |  |  |  |  |  |  |
| total |  | 16 (6 - 25) |  |  | 4 (1 - 19) | W = 523, p = .02, adolescents > adults |
| none/low group |  | 6 (4 - 9) |  |  | 2 (0 - 4) |  |
| high group |  | 29 (20 - 33) |  |  | 20 (11 - 31) |  |
| Mean daily dose<sup>g</sup> (mg/day, *mean (SD)*) |  |  |  |  |  |  |
| total |  | 25.3 (13.4) |  |  | 34.2 (24.9) | t(37) = -1.59, p = .12 |
| none/low group |  | 15.8 (9.2) |  |  | 13.9 (14.3) |  |
| high group |  | 36.0 (8.2) |  |  | 54.5 (13.8) |  |
| Medication use at FU (*yes/no*) |  | 17/12 |  |  | 4/15 |  |
| Stimulant-treatment naive at FU (*yes/no*) |  | 3/29 |  |  | 5/19 |  |
| ePOD-MPH RCT treatment group (*MPH/placebo*) |  | 11/21 |  |  | 11/13 |  |
| **Scan interval** |  |  |  |  |  |  |
| Time (months, *mean (SD)*) |  | 48.6 (14.2) |  |  | 53.3 (9.1) | t(54) = -1.41, p = .16 |

ADHD=attention-deficit/hyperactivity disorder, BL=baseline, FU=follow-up, IQ=intelligence quotient, MPH=methylphenidate.
<sup>a</sup> Paired samples t-test, two-sample t-test or Mann Whitney U test.
<sup>b</sup> For adolescents: subtest Wechsler Intelligence Scale for Children (WISC); for adults: National Adult Reading Test (NART, Dutch translation).
<sup>c</sup> Inattentive and hyperactive/impulsive subscales of the Disruptive Behavior Disorder Rating-Scale (DBD-RS).
<sup>d</sup> ADHD-Rating Scale (ADHD-RS) total score.
<sup>e</sup> For adolescents: Screen for Child Anxiety Related Disorders (SCARED); for adults: Beck Anxiety Inventory (BAI).
<sup>f</sup> For adolescents: Child Depression Inventory (CDI); for adults: Beck Depression Inventory (BDI).
<sup>g</sup> Converted to methylphenidate-equivalents.
<sup>h</sup> Calculated with a 30-day permissible gap.

### Stimulant medication use

Exposure duration was higher in adolescents than adults (W=509, P=0.04), whereas cumulative and mean daily dose of stimulant medication did not differ between age groups (Table 1, Figure 2). Two adult participants were prescribed the non-stimulant medication atomoxetine as treatment for ADHD, in addition to stimulant medication.

**Figure 2.**
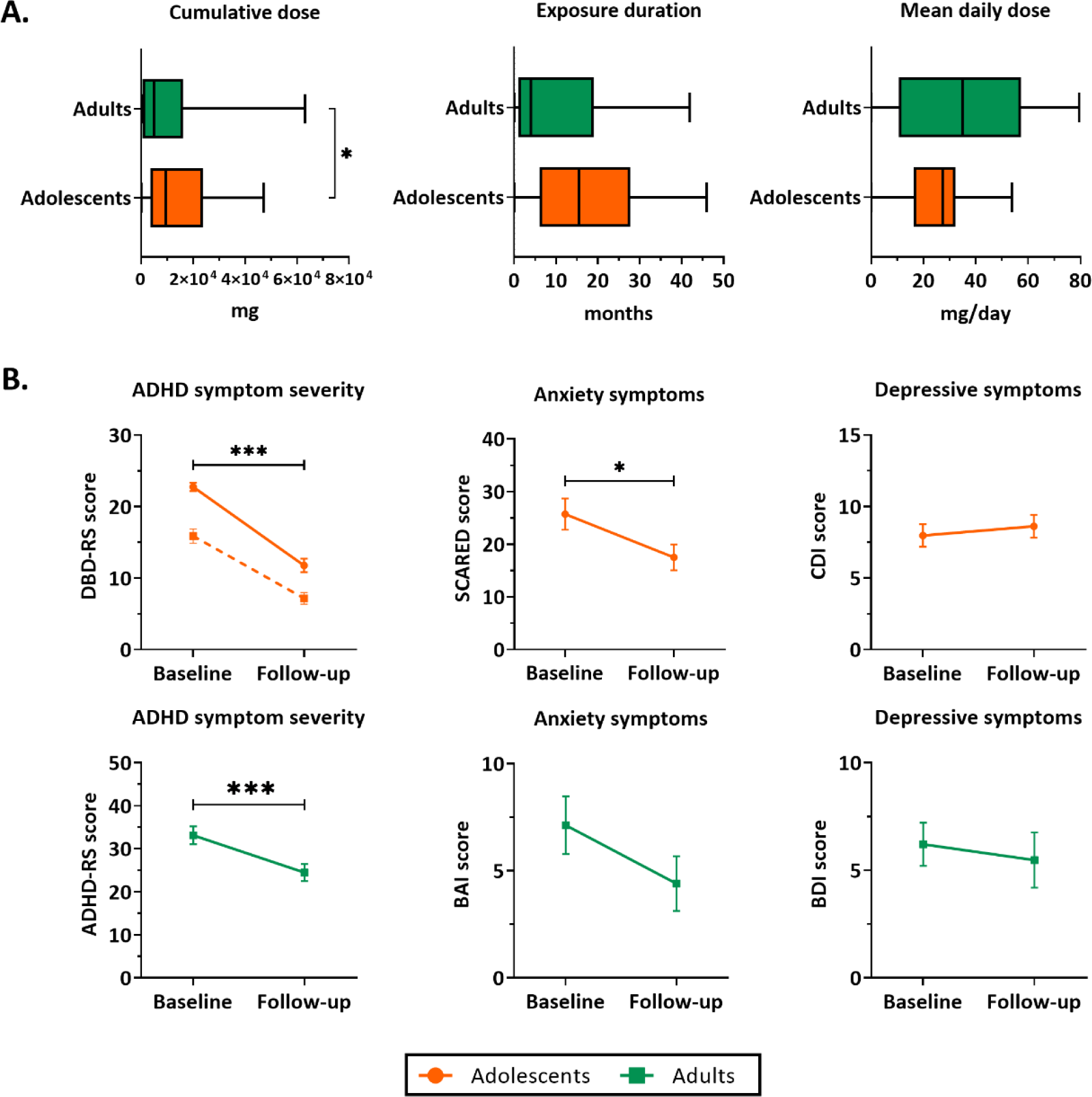
Stimulant medication use and clinical outcomes. A) Boxplots representing stimulant medication use (median and interquartile range) between baseline and 4-year follow-up assessment. B) ADHD symptom severity, anxiety and depressive symptom scores (mean ± SEM) at baseline and follow-up. For ADHD symptom severity in adolescents, the solid line represents inattentive symptoms and the dotted line represents hyperactive/impulsive symptoms. ADHD=attention-deficit/hyperactivity disorder, ADHD-RS=ADHD-Rating Scale, BAI=Beck Anxiety Inventory, BDI=Beck Depression Inventory, CDI=Child Depression Inventory, DBD-RS=Disruptive Behavioral Disorder Rating Scale (inattentive and hyperactive/impulsive subscales), SCARED=Screen for Child Anxiety Related Disorders. ***P<.001, *P<.05.

**Figure 3.**
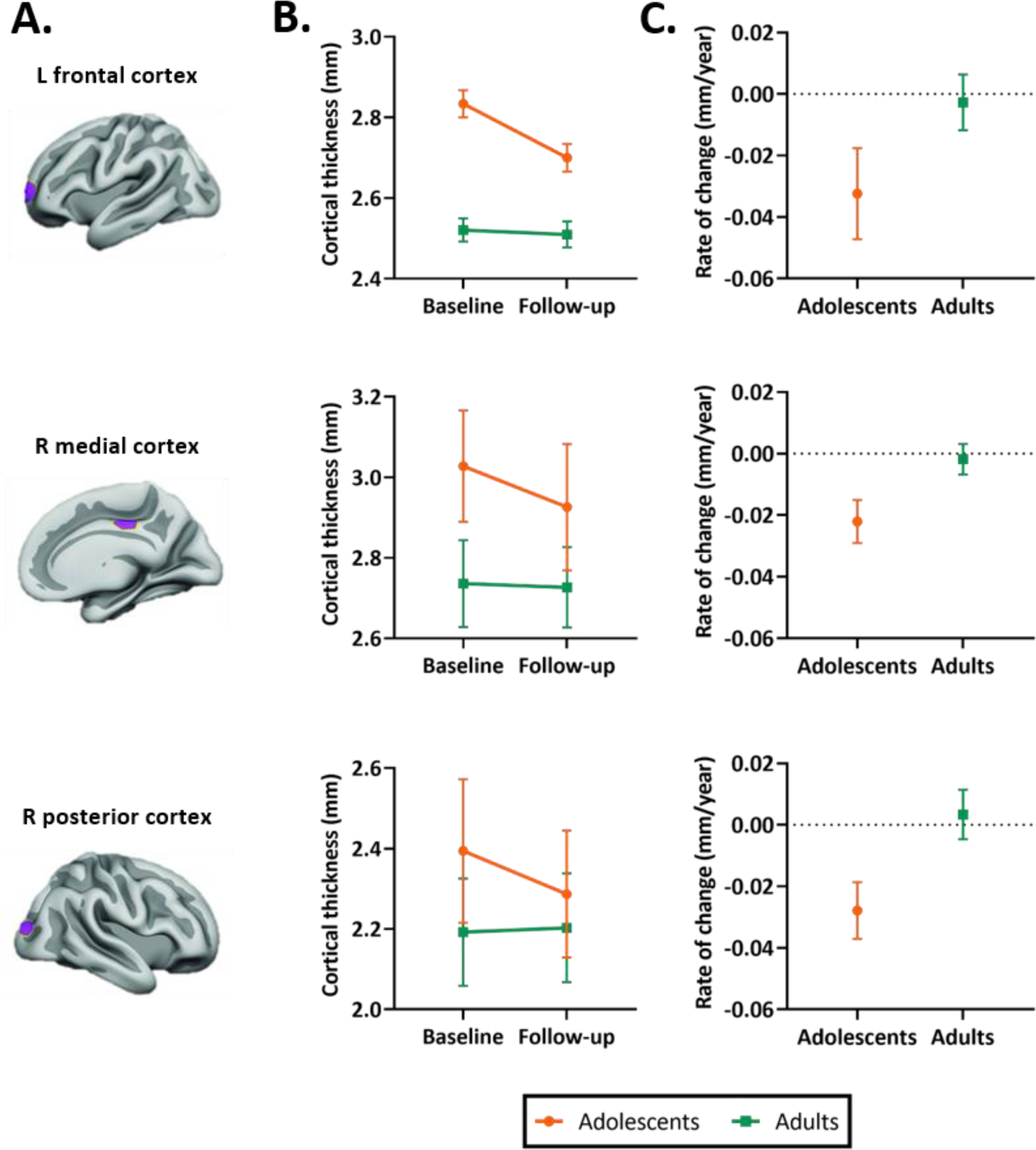
Selected regions of interest (ROIs) and apparent cortical thickness (mm) per ROI. A) Brain templates showing the selected ROIs in purple. B) Apparent cortical thickness (mean ± SEM) at baseline and 4-year follow-up assessment. Linear mixed models revealed an age-by-time interaction effect in all ROIs (P=.002 in left frontal ROI, P<.001 in right medial and posterior ROIs), reflecting apparent cortical thinning in adolescents but not adults. C) Rate of change in apparent cortical thickness (mean ± SEM) between baseline and 4-year follow-up assessment. The plotted values are the raw cortical thickness trajectories, without taking into account the covariates demeaned age at baseline and demeaned scan interval. ROI figures adapted from Walhovd et al (2020) [18].

### Clinical and behavioural outcomes

In both age groups, we found a reduction in ADHD symptom severity at 4-year follow-up compared to baseline. Symptoms of anxiety were reduced at follow-up in adolescents (t(28)=2.40, P=0.03), but not in adults (t(19)=1.51, P=0.15). Symptoms of depression did not significantly change between baseline and follow-up in either age group (adolescent: t(29)=-0.50, P=0.62; adults: t(20)=0.28, P=0.79) (Table 1, Figure 2).

In adults, we found an association between medication use and change in ADHD symptom scores (cumulative dose: r=-0.07, P=0.001; exposure duration: r=-0.73, P=0.001). No further associations were identified between medication use variables and baseline ADHD symptom severity, age at follow-up assessment, age at start of medication use, or change in weight between baseline and follow-up (Supplemental Table 1).

### ROI cortical thickness analysis

LMM analysis revealed no effects of cumulative dose and exposure duration on apparent cortical thickness development between baseline and follow-up of all ROIs, both for the models with continuous and grouped medication use variables, as well as when running the analysis split by age group (Table 2). All Bayes Factors were <1/100, providing strong evidence for no effects of cumulative dose and exposure duration on regional cortical thickness (Table 3). Furthermore, in adults, demeaned age at baseline had an effect on apparent cortical thickness of the right medial ROI (cumulative dose: t(24)=-2.36, P=0.03; exposure duration: t(24)=-2.39, P=0.03), indicating that higher age at baseline was associated with lower apparent cortical thickness of the right medial ROI. At 4-year follow-up, we found no effects of ePOD-MPH RCT treatment groups (MPH, placebo) on apparent cortical thickness of all ROIs (Table 2).

**Table 2.** Statistics of the confirmatory region of interest (ROI) analysis.

| Predictors | Left Prefrontal ROI |  |  | Right Medial ROI |  |  | Right Posterior Parietal ROI |  |  |
| --- | --- | --- | --- | --- | --- | --- | --- | --- | --- |
|  | Beta | CI | p-value | Beta | CI | p-value | Beta | CI | p-value |
| *Cumulative dose (CD, continuous)* |  |  |  |  |  |  |  |  |  |
| CD | -0.00 | -0.00 – 0.00 | .95 | -0.00 | -0.00 – 0.00 | .98 | 0.00 | -.00 – .00 | .98 |
| Time | -0.12 | -0.19 – -0.05 | **.002** | -0.09 | -0.13 – -0.05 | **<.001** | -0.10 | -.16 – -.05 | **<.001** |
| Age | -0.30 | -0.42 – -0.17 | **<.001** | -0.27 | -0.36 – -0.17 | **<.001** | -0.22 | -.33 – -.11 | **<.001** |
| CD * Time | -0.00 | -0.00 – 0.00 | .72 | -0.00 | -0.00 – 0.00 | .60 | -0.00 | -.00 – .00 | .58 |
| CD * Age | -0.00 | -0.00 – 0.00 | .74 | -0.00 | -0.00 – 0.00 | .57 | 0.00 | -.00 – .00 | .79 |
| Time * Age | 0.12 | 0.02 – 0.22 | **.02** | 0.08 | 0.03 – 0.13 | **.002** | 0.11 | .04 – .18 | **.003** |
| CD * Time * Age | -0.00 | -0.00 – 0.00 | .91 | 0.00 | -0.00 – 0.00 | .70 | 0.00 | -.00 – .00 | .64 |
| Demeaned age at BL | -0.01 | -0.02 – 0.01 | .33 | -0.01 | -0.02 – 0.00 | .16 | 0.00 | -.01 – .02 | .62 |
| Standardised scan interval | 0.00 | -0.00 – 0.00 | .75 | -0.00 | -0.00 – 0.00 | .35 | 0.00 | -.00 – .00 | .50 |
| *Exposure duration (ED, continuous)* |  |  |  |  |  |  |  |  |  |
| ED | -0.00 | -0.01 – 0.00 | .85 | 0.00 | -0.00 – 0.00 | .98 | -0.00 | -0.00 – 0.00 | .98 |
| Time | -0.13 | -0.21 – -0.05 | **.002** | -0.09 | -0.13 – -0.05 | **<.001** | -0.10 | -0.16 – -0.04 | **.001** |
| Age | -0.29 | -0.42 – -0.16 | **<.001** | -0.26 | -0.36 – -0.16 | **<.001** | -0.20 | -0.32 – -0.09 | **.001** |
| ED * Time | 0.00 | -0.00 – 0.00 | >.99 | -0.00 | -0.00 – 0.00 | .57 | -0.00 | -0.00 – 0.00 | .62 |
| ED * Age | -0.00 | -0.01 – 0.00 | .48 | -0.00 | -0.01 – 0.00 | .38 | -0.00 | -0.01 – 0.01 | .90 |
| Time * Age | 0.14 | 0.04 – 0.24 | **.01** | 0.08 | 0.03 – 0.13 | **.004** | 0.11 | 0.04 – 0.19 | **.004** |
| ED * Time * Age | -0.00 | -0.01 – 0.00 | .49 | 0.00 | -0.00 – 0.00 | .78 | 0.00 | -0.00 – 0.00 | .80 |
| Demeaned age at BL | -0.01 | -0.02 – 0.01 | .29 | -0.01 | -0.02 – 0.00 | .16 | 0.00 | -0.01 – 0.01 | .75 |
| Standardised scan interval | 0.00 | -0.00 – 0.00 | .74 | -0.00 | -0.00 – 0.00 | .35 | 0.00 | -0.00 – 0.00 | .49 |
| *Cumulative dose (CD, grouped)\** |  |  |  |  |  |  |  |  |  |
| CD | 0.02 | -0.10 – 0.14 | .71 | -0.00 | -0.09 – 0.09 | .96 | 0.03 | -0.08 – 0.14 | .57 |
| Time | -0.11 | -0.17 – -0.04 | **.002** | -0.09 | -0.12 – -0.05 | **<.001** | -0.10 | -0.14 – -0.05 | **<.001** |
| Age | -0.26 | -0.40 – -0.13 | **<.001** | -0.28 | -0.38 – -0.18 | **<.001** | -0.19 | -0.31 – -0.07 | **.003** |
| CD * Time | -0.05 | -0.14 – 0.05 | .34 | -0.02 | -0.07 – 0.02 | .34 | -0.04 | -0.11 – 0.03 | .31 |
| CD * Age | -0.09 | -0.29 – 0.10 | .35 | -0.01 | -0.15 – 0.14 | .91 | -0.04 | -0.21 – 0.14 | .68 |
| Time * Age | 0.10 | 0.00 – 0.20 | **.046** | 0.09 | 0.04 – 0.14 | **<.001** | 0.11 | 0.04 – 0.18 | **.005** |
| CD * Time * Age | 0.03 | -0.11 – 0.17 | .68 | -0.01 | -0.08 – 0.06 | .71 | 0.03 | -0.07 – 0.14 | .53 |
| Demeaned age at BL | -0.01 | -0.02 – 0.01 | .24 | -0.01 | -0.02 – 0.00 | .21 | 0.00 | -0.01 – 0.02 | .80 |
| Standardised scan interval | 0.00 | -0.00 – 0.00 | .74 | -0.00 | -0.00 – 0.00 | .39 | 0.00 | -0.00 – 0.00 | .54 |
| *Exposure duration (ED, grouped)\** |  |  |  |  |  |  |  |  |  |
| ED | -0.01 | -0.13 – 0.11 | .88 | 0.00 | -0.08 – 0.09 | .91 | -0.00 | -0.11 – 0.11 | .98 |
| Time | -0.12 | -0.19 – -0.06 | **<.001** | -0.09 | -0.12 – -0.06 | **<.001** | -0.13 | -0.17 – -0.08 | **<.001** |
| Age | -0.32 | -0.44 – -0.19 | **<.001** | -0.28 | -0.37 – -0.19 | **<.001** | -0.19 | -0.30 – -0.09 | **.001** |
| ED * Time | -0.02 | -0.11 – 0.08 | .73 | -0.01 | -0.06 – 0.03 | .55 | 0.03 | -0.04 – 0.10 | .44 |
| ED * Age | 0.01 | -0.18 – 0.20 | .92 | -0.01 | -0.15 – 0.13 | .87 | -0.03 | -0.20 – 0.14 | .72 |
| Time * Age | 0.13 | 0.04 – 0.23 | **.008** | 0.09 | 0.04 – 0.14 | **.001** | 0.14 | 0.07 – 0.21 | **<.001** |
| ED * Time * Age | -0.03 | -0.18 – 0.11 | .65 | -0.01 | -0.08 – 0.07 | .87 | -0.04 | -0.14 – 0.07 | .51 |
| Demeaned age at BL | -0.01 | -0.02 – 0.01 | .41 | -0.01 | -0.02 – 0.00 | .22 | 0.00 | -0.01 – 0.01 | .87 |
| Standardised scan interval | 0.00 | -0.00 – 0.00 | .75 | -0.00 | -0.00 – 0.00 | .34 | 0.00 | -0.00 – 0.00 | .54 |
| *ePOD-MPH RCT treatment group* |  |  |  |  |  |  |  |  |  |
| Treatment# | 0.01 | -0.11 – 0.13 | .88 | 0.01 | -0.09 – 0.10 | .90 | 0.06 | -0.05 – 0.17 | .32 |
| Time | -0.13 | -0.21 – -0.05 | **.003** | -0.12 | -0.16 – -0.07 | **<.001** | -0.10 | -0.16 – -0.04 | **.003** |
| Age | -0.39 | -0.53 – -0.25 | **<.001** | -0.26 | -0.36 – -0.15 | **<.001** | -0.20 | -0.33 – -0.07 | **.003** |
| Treatment# * Time | -0.00 | -0.10 – 0.10 | .99 | 0.03 | -0.02 – 0.08 | .28 | -0.03 | -0.10 – 0.05 | .49 |
| Treatment# * Age | 0.15 | -0.03 – 0.33 | .11 | -0.05 | -0.19 – 0.09 | .48 | -0.00 | -0.17 – 0.16 | .98 |
| Time * Age | 0.15 | 0.04 – 0.26 | **.01** | 0.10 | 0.04 – 0.15 | **.002** | 0.10 | 0.01 – 0.18 | **.03** |
| Treatment# * Time * Age | -0.06 | -0.20 – 0.09 | .45 | -0.01 | -0.08 – 0.07 | .84 | 0.04 | -0.06 – 0.15 | .42 |
| Demeaned age at BL | -0.01 | -0.02 – 0.00 | .22 | -0.01 | -0.02 – 0.00 | .31 | 0.00 | -0.01 – 0.01 | .88 |
| Standardised scan interval | 0.00 | -0.00 – 0.00 | .90 | -0.00 | -0.00 – 0.00 | .36 | 0.00 | -0.00 – 0.00 | .55 |

BL = baseline, CI = 95% confidence interval, RCT = randomised controlled trial, ROI = Region of interest.
\* Median split.

**Table 3.** Bayes Factors comparing the models with (continuous) medication use to the models without medication use.

| **Medication use variable** | **Region of interest** | **Bayes Factor** | **Interpretation** |
| --- | --- | --- | --- |
| cumulative dose | Left prefrontal | $1.8 * 10^{-04}$ | Extreme evidence for null Hypothesis |
| cumulative dose | Right Medial | $1.6 * 10^{-04}$ | Extreme evidence for null Hypothesis |
| cumulative dose | Right Posterior Parietal | $1.1 * 10^{-04}$ | Extreme evidence for null Hypothesis |
| exposure duration | Left prefrontal | $4.8 * 10^{-04}$ | Extreme evidence for null Hypothesis |
| exposure duration | Right Medial | $2.1 * 10^{-04}$ | Extreme evidence for null Hypothesis |
| exposure duration | Right Posterior Parietal | $9.8 * 10^{-05}$ | Extreme evidence for null Hypothesis |

In all ROIs, we identified an age-by-time interaction effect on apparent cortical thickness (left prefrontal ROI: t(55)=3.26, P=0.002; right medial ROI: t(54)=4.75, P<0.001; right posterior parietal ROI: t(54)=4.68, P<0.001) (Figure 3B). Post hoc analyses revealed a decrease in cortical thickness of all ROIs between baseline and 4-year follow-up in adolescents (left prefrontal ROI: t(31)=-4.53, P<0.001; right medial ROI: t(30)=-6.74, P<0.001; right posterior parietal ROI: t(30)=-5.75, P<0.001), but not in adults (left prefrontal ROI: t(24)=0.14, P=0.89; right medial ROI: t(24)=-0.97, P=0.34; right posterior parietal ROI: t(24)=0.68, P=0.50). Figure 3C presents the rate of change (mm/year) for each ROI (Supplemental Results).

## Discussion

This study investigated the age-dependency of long-term stimulant-treatment effects on regional apparent cortical thickness in adolescents and adults previously diagnosed with ADHD. In contrast to the 4-month ePOD-MPH RCT, in this 4-year naturalistic follow-up we observed no evidence for long-term effects of stimulant medication use on apparent cortical thickness in any of the 3 ROIs investigated (left prefrontal, right medial and right posterior parietal ROIs). Moreover, as hypothesised, the treatment conditions from the ePOD-MPH RCT could no longer be distinguished. We did, however, identify differences in apparent cortical thickness development between adolescents and adults, which were consistent with existing literature on typical cortical development [7,30]. In addition, we observed improvements in clinical outcomes, as well as an association between higher medication use and lower change in ADHD symptoms in adults. A possible explanation for this association may be that participants with greater improvement in ADHD symptom severity may no longer require stimulant medication.

In contrast to our hypothesis and prior short-term findings [7], we observed no evidence for long-term stimulant-treatment effects on (regional) cortical thickness development, either in adolescents or adults. This finding is in contrast with the neurochemical imprinting hypothesis, which states that stimulant administration during development may have lasting effects [17,18]. We propose several explanations for our findings. First, the short-term effects of stimulant medication use identified in the initial ePOD-MPH RCT may be transient, supporting the neural plasticity hypothesis that the (human) brain is able to adapt in response to internal and external stimuli [31]. Alternatively, the current study may have been unable to detect subtle stimulant-treatment effects due to limited sample size related to loss-to-follow-up of participants, as well as heterogeneous medication use and prescription adherence among participants [32,33]. Nonetheless, the Bayes Factors calculated here provide strong evidence for the absence of stimulant medication effects on apparent cortical thickness development. Moreover, previous large-scale multicentre projects with heterogeneous study populations also reported no evidence for stimulant-treatment effects on cortical thickness [15,16].

Discrepant findings in literature regarding stimulant-treatment effects on apparent cortical thickness in ADHD can be attributed to various reasons. Firstly, differences in age of the study participants may influence findings, as the cerebral cortex continues to develop throughout childhood and adolescence into adulthood. As a result, assessment of stimulant medication effects in different developmental stages may yield different findings. Furthermore, previous studies used differing approaches to calculate stimulant medication use or treatment profiles [13,15,16]. Findings may also be impacted by methodological decisions, such as use of an ROI or whole-brain approach, MR field strength and scanning parameters, or cortical thickness estimation technique [19,12].

A previous study observed that the mean rate of apparent cortical thinning in stimulant-treated ADHD participants was comparable to typically developing peers, while ADHD participants *off* stimulant medication showed more rapid apparent cortical thinning [13]. Similarly, in our study, we observed comparable changes in apparent cortical thickness development during the naturalistic follow-up. However, we need to be cautious with speculations about associations of medication use with ‘normalisation’ of cortical thickness development, since only few participants in our sample remained stimulant treatment-naïve and no typically developing control group was included. To gain further insights, it is essential to conduct large-scale longitudinal studies that include stimulant-treated and untreated individuals with ADHD, as well as typically developing peers.

A critical strength of this study is its longitudinal design with stimulant treatment-naive participants at baseline, ruling out the influence of a history of medication use on cortical development. Moreover, we replicated previous findings of age-dependent apparent cortical thickness development [6,30] and rate of regional cortical thinning in stimulant-treated adolescents with ADHD [13]. Some limitations should also be considered. First, analogous to previous naturalistic studies, we assumed similar prescription adherence (complete adherence) for all participants. Nevertheless, medication adherence rates have been found to vary substantially [32,33], therefore future studies should consider using reliable treatment adherence measures [34]. Other limitations are the restricted sample size at follow-up assessment and the use of different MR scanners at baseline and follow-up. However, we were still able to identify general neurodevelopmental patterns of relatively faster reductions in apparent cortical thickness in adolescents than in adults, which aligns with existing literature [6,30]. Furthermore, the results of this study cannot be extrapolated to all adolescents and adults with ADHD, since only male participants within a specific age range were included. This choice was based on the knowledge that patterns of brain development differ considerably between males and females [35], and that the prevalence of ADHD is higher in males than in females [2].

In conclusion, this study found no evidence for long-term effects of stimulant-treatment on apparent cortical thickness development in adolescents and adults previously diagnosed with ADHD. Future research should include prescription adherence measures and employ standardised/homogeneous approaches for acquisition and analysis. Moreover, there is a need for longitudinal studies including stimulant-treated and untreated individuals with ADHD, as well as typically developing peers, to improve our understanding of (age-dependent) effects of stimulant-treatment on the developing brain.

## Data Availability

All data produced in the present study are available upon reasonable request to the authors.

## Abbreviations

ADHD: attention-deficit/hyperactivity disorder
ePOD-MPH: effects of Psychotropic drugs On the Developing brain - methylphenidate
MPH: methylphenidate
RCT: randomised controlled trial
ROI: region of interest

## Acknowledgements

We would like to thank all participants and their parents for their contribution to this study and all students that helped with collection and analysis of the data.

