## Supplemental Material for "Stimulant medication use and apparent cortical thickness development in attention-deficit/hyperactivity disorder: a prospective longitudinal study"

### Supplemental Materials

#### SUPPLEMENTAL METHODS

##### Participants and recruitment

For the initial randomised controlled trial of the "effects of Psychotropic drugs On the Developing brain - methylphenidate" project (ePOD-MPH RCT), a total of 50 children (10-12 years of age) and 49 adult (23-30 years of age) male outpatients diagnosed with attention-deficit/hyperactivity disorder (ADHD; all subtypes) and in need of pharmacological therapy were included, as described elsewhere [1]. Participants were recruited through clinical programs at the Department of Child and Adolescent Psychiatry at Triversum (Alkmaar, the Netherlands), De Bascule Academic Center for Child and Adolescent Psychiatry (Amsterdam, the Netherlands), and PsyQ mental health facility (The Hague, the Netherlands).

Exclusion criteria were: comorbid axis I psychiatric disorders requiring treatment with medication at study entry, estimated IQ < 80 (assessed using a subtest of the Wechsler Intelligence Scale for children-revised (WISC-III-R [2]) in children, and the National Adult Reading Test (NART [3]) in adults), a history of major neurological or medical illness or clinical treatment with drugs influencing the dopamine system (for adults before 23 years of age), such as stimulants, neuroleptics, antipsychotics, and/or D2/3 agonists (see [4] for more detail).

##### Stimulant medication use

During the ePOD-MPH RCT participants received oral dosages of short-acting MPH, starting with 1–2 doses of 0.3 mg/kg daily. Dosages were increased weekly with 5–10 mg/day to a maximum of 50 mg/day until the target clinical dosage was reached, in line with clinical guidelines in the

Netherlands. If, after in- or decreasing the dosage, serious side-effects occurred, the participant returned to the previous dosage and dosage modifications were more gradual thereafter. Cumulative dose of MPH was calculated from the prescribed medication and the treatment compliance rate. For one subject, compliance rate during the trial was unknown and a 100% compliance rate was assumed.

Cumulative dose after the ePOD-MPH RCT was calculated by multiplying the number of days of medication use for each prescription (prescription start date subtracted from the prescription end date) with the prescribed daily dose (assuming complete adherence). If the prescription start and end date were not available, the number of prescribed units was used. Exposure duration to stimulant medication was determined by calculating the time between the start date and end date of stimulant medication use, with a 30-day permissible gap to allow for commonly occurring "medication holidays", and rounding to months. Next, mean daily dose between baseline and 4-year follow-up assessment was calculated by dividing cumulative dose (in mg) by exposure duration (in days). The age at start of medication use was determined based on the date of the baseline assessment for participants who received MPH during the ePOD-MPH RCT, and date of first stimulant medication prescription in the pharmacy overview for participants who received placebo during the ePOD-MPH RCT.

Participants with no stimulant medication use between baseline and 4-year follow-up assessment were considered stimulant treatment-naive, and cumulative dose, exposure duration, and mean daily dose were set to zero.

**Region of interest definition**

The Montreal Neurological Institute (MNI) coordinates for previously [5] reported peak group differences were as follows: left frontal: −26, 61, 6; right medial frontal: 6, −23, 40; right posterior:

30, −93, 12. MNI coordinates of the vertices corresponding to these peak group differences were converted to the Talairach coordinate system, and dilated 15 times to create hexagonal regions of interest (ROIs) of 550 $mm^2$ when transformed to subjects' brain surfaces.

**Bayes Factor calculation and interpretation**

The bayesfactor_models() function of the bayestestR package (version 0.9.0) was used for Bayes Factor calculation, comparing the models with medication use to the models without medication use. The full models with medication use assessed the main and interaction effects of stimulant medication use (cumulative dose, exposure duration), time (baseline, follow-up) and age group (adolescents, adults) on regional apparent cortical thickness. The null models without medication use assessed the main and interaction effects of time (baseline, follow-up) and age group (adolescents, adults) on regional apparent cortical thickness. For interpretation of the calculated Bayes Factors, please see Supplemental Table 1.

**Exploratory analyses**

Linear mixed models (LMMs) were constructed separately per age group to explore main effects of main effects and interactions of clinical outcome measures and time (baseline, follow-up) on regional apparent cortical thickness. Separate models were used for each ROI and clinical outcome measure (ADHD symptom severity, anxiety and depressive symptoms). In line with the ROI cortical thickness analysis, demeaned age at baseline and standardised scan interval were included in the models as covariates. Benjamini-Hochberg multiple comparison correction (FDR=5%) was applied to adjust for the three clinical scales evaluated.

Next, we performed an exploratory whole-brain analysis in Freesurfer to assess associations between medication use and vertex-wise cortical thickness and surface area, separately for adolescents and adults. Again, demeaned age at baseline and standardised scan interval were included in the models as covariates. Separate models were used for each surface measure (cortical thickness, surface area), and medication use variable (mean daily dose, medication use at time of 4-year follow-up). First, each individual's surface was sampled to the fsaverage surface and the difference between the two time points (baseline, follow-up) was computed. Next, these differences were concatenated into one file and smoothing was performed (fwhm 10). Subsequently, paired analysis was performed using mri_glmfit, and cluster correction was performed using mri_glmfit-sim (cluster threshold: P<.001, FDR=5%) to account for the amount of smoothing and the number of contiguous significant vertices.

**Deviations from pre-registered analysis plan**

Some deviations were made from the pre-registered analysis plan:

1. Medication use variables were grouped using a median split per age group, rather than across age groups.
2. The medication use variables were not transformed, as the residuals of the LMMs exhibited a normal distribution. Additionally, transformation of the medication use variables did not lead to an improvement in the distribution of residuals or model fit.
3. To account for the differences in clinical measures between adolescents and adults, LMMs with clinical outcomes were constructed separately for adolescents and adults. This decision was made because different scales were used to assess clinical measures in these two age groups, rendering the symptom scores incomparable across them.
4. Associations between ADHD symptom severity and apparent cortical thickness were also assessed, although this was not initially included in the pre-registered analysis plan. This omission was an oversight in the planning process.
5. The change in ADHD symptom severity was associated with medication use (cumulative dose and exposure duration). Therefore, ADHD symptom severity was not included as a covariate in the LMMs.

6. For the exploratory whole-brain analysis, we evaluated associations between stimulant medication and change in apparent cortical thickness and surface area separately for adolescents and adults, rather than using LMMs to assess the main effects and interactions of medication use, age group and time. This decision was made to facilitate interpretation and reduce the need for multiple testing. Moreover, multiple comparison correction (FDR=5%) was performed using the cluster correction function provided by Freesurfer, rather than using the Benjamini-Hochberg method.

#### SUPPLEMENTAL RESULTS

##### Sensitivity analysis without participants with structural abnormalities

Our findings were robust when excluding two adolescents with structural abnormalities from the sample. The only difference with the main findings was that we found a main effect of exposure duration ($t(54)=-2.52$, $p=.02$) on apparent cortical thickness of the left prefrontal ROI, although this was only identified after removal of all two-way and three-way interaction effects involving medication use. Moreover, the corresponding Bayes Factor was <1/100, providing strong evidence for no effects of exposure duration on apparent cortical thickness of this ROI.

##### Exploratory analyses

*Relations between regional apparent cortical thickness and clinical outcomes*

For clinical outcomes, in adolescents, after correction for multiple comparisons we identified a depression score-by-time interaction effect on cortical thickness of the right posterior parietal ROI ($t(32) = -2.99$, adjusted $P = .008$). In adults, we identified ADHD symptom score-by-time ($t(20) = 2.36$, adjusted $P = .04$) and anxiety score-by-time ($t(21) = 3.00$, adjusted $P = .005$) interaction effects on cortical thickness of the left prefrontal ROI. Post hoc analysis revealed opposite correlations between apparent cortical thickness and clinical scores at baseline compared with 4-year follow-up, although none of these associations were significant (Supplemental Table 4). We found no other main or interaction effects of clinical outcomes on apparent cortical thickness in adolescents or adults.

*Whole-brain analysis of apparent cortical thickness and surface area*

In both age groups, we identified no additional associations between medication use and apparent cortical thickness or surface area.

**Supplemental Table 1. Thresholds for interpretation of Bayes Factors.**

| Bayes Factor | Interpretation |
|---|---|
| 1 | No evidence for null Hypothesis |
| 1/3 | Anecdotal evidence for null Hypothesis |
| 1/3 - 1/10 | Moderate evidence for null Hypothesis |
| 1/10 - 1/30 | Strong evidence for null Hypothesis |
| 1/30 - 1/100 | Very strong evidence for null Hypothesis |
| < 1/100 | Extreme evidence for null Hypothesis |

**Supplemental Table 2. Baseline characteristics of participants that did and did not participate in the 4-year follow-up assessment.** Data are presented as mean (standard deviation) or fraction (MPH/placebo). Comparisons were made prior to data processing and analysis.

| | **Adolescents** | | | **Adults** | | |
|---|---|---|---|---|---|---|
| | Return = yes<br>n = 33 | Return = no<br>n = 17 | Statistics[a] | Return = yes<br>n = 25 | Return = no<br>n = 24 | Statistics[a] |
| Age<br>(years, *mean (SD)*) | 11.2 (0.9) | 11.6 (0.8) | t(48) = 1.49, P = .14 | 29.8 (5.0) | 27.3 (3.8) | t(47) = -1.98, P = .053 |
| IQ[b]<br>*(mean (SD))* | 105.1 (17.7) | 102.3 (19.6) | t(46) = -0.50, P = .62 | 106.7 (4.9) | 109.0 (9.5) | t(43) = 1.05, P = .30 |
| ADHD-inattentive symptom severity[c]<br>*(mean (SD))* | 22.60 (3.43) | 21.31 (2.98) | t(47) = -1.29, P = .20 | | | |
| ADHD-hyperactive/impulsive symptom severity[c]<br>*(mean (SD))* | 15.85 (5.51) | 14.81 (6.50) | t(47) = -0.58, P = .56 | | | |
| ADHD symptom severity[d]<br>*(mean (SD))* | | | | 33.00 (9.71) | 32.38 (9.83) | t(43) = -0.21, P = .83 |
| Anxiety symptoms[e]<br>*(mean (SD))* | 26.03<br>(16.58) | 30.71<br>(17.50) | t(48) = 0.93, P = .36 | 6.84 (6.61) | 10.64 (7.03) | t(45) = 1.91, P = .06 |
| Depressive symptoms[f]<br>*(mean (SD))* | 8.31 (4.75) | 8.29 (3.85) | t(47) = -0.01, P = .99 | 6.21 (4.90) | 8.18 (6.68) | t(44) = 1.15, P= .26 |
| ePOD-MPH RCT treatment group<br>(*MPH/placebo)* | 12/21 | 13/4 | $X^2(1) = 5.70$, P = .02 | 12/13 | 13/11 | $X^2(1) = 0.02$, P = .88 |

ADHD=attention-deficit/hyperactivity disorder, IQ=intelligence quotient, MPH=methylphenidate.
[a] Two-sample t-test or Chi-squared test.
[b] For adolescents: subtest Wechsler Intelligence Scale for Children (WISC); for adults: National Adult Reading Test (NART, Dutch translation).
[c] Inattentive and hyperactive/impulsive subscales of the Disruptive Behavior Disorder Rating-Scale (DBD-RS).
[d] ADHD-Rating Scale (ADHD-RS) total score.
[e] For adolescents: Screen for Child Anxiety Related Disorders (SCARED); for adults: Beck Anxiety Inventory (BAI).
[f] For adolescents: Child Depression Inventory (CDI); for adults: Beck Depression Inventory (BDI).

**Supplemental Table 3. Correlations between medication use and other/clinical variables.** Spearman correlations were used, unless indicated otherwise.

| | **Cumulative dose (mg)** | **Exposure duration (months)** | **Mean daily dose (mm/day)** |
|---|---|---|---|
| **Adolescents** | | | |
| BL ADHD symptoms (inattentive)[b] | r = .02, P = .44 | r = .08, P = .66 | r = .06, P = .77 |
| Change in ADHD symptoms (inattentive)[b] | r = -.24, P = .22 | r = -.18, P = .36 | r = -.16, P = .44 |
| BL ADHD symptoms (hyperactive/impulsive)[b] | r = .35, P = .06 | r = .26, P = .17 | r = .18, P = .35 |
| Change in ADHD symptoms (hyperactive/impulsive)[c] | r = -.39, P = .04 | r = -.31, P = .12 | r = -.33, P = .09 |
| Change in weight (*kg*) | r = .09, P = .66 | r = .11, P = .59 | r = .18, P = .37 |
| Age at FU (*years*) | r = .09, P = .63 | r = -.01, P = .96 | r = .34, P = .07 |
| Age at start medication use (*years*) | r = .08, P = .69 | r = -.08, P = .69 | r = .32, P = .09 |
| **Adults** | | | |
| BL ADHD symptoms[d] | r =.10, P = .70 | r = .20, P = .42 | r = -.39, P = .14[a] |
| Change in ADHD symptoms[d] | r = -.07, P = .0014* | r = -.73, P = .0014* | r = -.18, P = .46 |
| Change in weight (*kg*) | r = -.28, P = .24 | r = -.29, P = .22 | r = -.43, P = .07[a] |
| Age at FU (*years*) | r = -.11, P = .67 | r = .01, P = .97 | r = -.10, P = .07 |
| Age at start medication use (*years*) | r = -.23, P = .34 | r = -.11, P = .65 | r = -.33, P = .16[a] |

ADHD=attention-deficit/hyperactivity disorder, BL=baseline, FU=4-year follow-up.
[a] Spearman correlation coefficient.
[b] Inattentive and hyperactive/impulsive subscales of the Disruptive Behavior Disorder Rating Scale (DBD-RS).
[c] ADHD-rating scale (ADHD-RS) total score.
*Significant after Benjamini-Hochberg multiple comparison correction (FDR=5%; adolescents: 7 tests, adults: 5 tests).

**Supplemental Table 4**. Mean rate of change in apparent cortical thickness (mm/year) during the ePOD-MPH RCT and 4-year follow-up.

| | **Left Prefrontal** *mm/year* | **Right Medial** *mm/year* | **Right Posterior Parietal** *mm/year* |
|---|---|---|---|
| ePOD-MPH RCT* | | | |
| **Adolescent** | | | |
| *MPH* | -0.133 ± 0.572 | 0.048 ± 0.252 | -0.019 ± 0.363 |
| *PLC* | -0.066 ± 0.748 | -0.132 ± 0.290 | -0.094 ± 0.403 |
| **Adult** | | | |
| *MPH* | 0.124 ± 0.385 | -0.033 ± 0.193 | 0.044 ± 0.191 |
| *PLC* | -0.028 ± 0.326 | -0.002 ± 0.179 | 0.042 ± 0.153 |
| ePOD-MPH FU | | | |
| **Adolescent** | -0.032 ± 0.040 | -0.022 ± 0.019 | -0.028 ± 0.025 |
| **Adult** | -0.003 ± 0.021 | -0.002 ± 0.012 | 0.003 ± 0.019 |

*During the ePOD-MPH RCT, mean rate of change was converted from the duration of the RCT to years.

**Supplemental table 5. Associations between clinical outcomes and regional apparent cortical thickness at baseline and 4-year follow-up assessment.** Data are presented as Pearson correlation coefficients.

| | **Baseline** | **Follow-up** | **ROI** |
|---|---|---|---|
| **Adolescent** | | | |
| Depression score*time | r = .16, P = .40 | r = -.24, P = .20 | Right posterior parietal |
| **Adult** | | | |
| ADHD symptom score*time | r = -.40, P = .06 | r = .20, P = .40 | Left prefrontal |
| Anxiety score*time | r = -.01, P = .97 | r = .07, P = .78 | Left prefrontal |

ADHD = attention-deficit/hyperactivity disorder, ROI = region of interest.
